# Creating an automated tool for a consistent and repeatable evaluation of disability progression in clinical studies for Multiple Sclerosis

**DOI:** 10.1101/2024.01.30.24302013

**Authors:** Noemi Montobbio, Luca Carmisciano, Alessio Signori, Marta Ponzano, Irene Schiavetti, Francesca Bovis, Maria Pia Sormani

**Affiliations:** Department of Health Sciences (DISSAL), University of Genova, Genova, Italy; IRCCS Ospedale Policlinico San Martino, Genoa, Italy

**Keywords:** multiple sclerosis, outcome assessment, clinical trial, observational study, software tools, Reproducibility of Finding

## Abstract

**Background:** The lack of standardized disability progression evaluation in multiple sclerosis (MS) hinders reproducibility of clinical study results, due to heterogeneous and poorly reported criteria.

**Objectives:** To demonstrate the impact of using different parameters when evaluating MS progression, and to introduce an automated tool for reproducible outcome computation.

**Methods:** Re-analyzing BRAVO clinical trial data (NCT00605215), we examined the fluctuations in computed treatment effect on confirmed disability progression (CDP) and progression independent of relapse activity (PIRA) when varying different parameters. These analyses were conducted using the *msprog* package for R, which we developed as a tool for CDP assessment from longitudinal data, given a set of criteria that can be specified by the user.

**Results:** The BRAVO study reported a hazard ratio (HR) of 0.69 (95%CI:0.46-1.02) for CDP. Using the different parameter configurations, the resulting treatment effect on CDP varied considerably, with HRs ranging from 0.59 (95%CI:0.41-0.86) to 0.72 (95%CI:0.48-1.07). The treatment effect on PIRA varied from an HR=0.62 (95%CI:0.41-0.93) to an HR=0.65 (95%CI:0.40-1.04).

**Conclusions:** The adoption of an open-access tool validated by the research community, with clear parameter specification and standardized output, could greatly reduce heterogeneity in CDP estimation and promote repeatability of study results.

## Introduction

In its chronic, neurodegenerative course, multiple sclerosis (MS) affects the central nervous system (CNS), resulting in gradual and irreversible disability progression. The Expanded Disability Status Scale (EDSS) is commonly used to assess and score such progression, serving as the primary outcome approved by regulators for clinical trials evaluating disease-modifying therapies in MS^1-2^. Despite its widespread use, the EDSS has well-documented and extensively studied inherent limitations^3^. It is based on the standard neurological examination, which is inherently subjective and susceptible to large variations in assessment^4^; it is highly weighted toward ambulation, while it captures poorly MS related cognitive and upper limb impairment; the scale used is a non-linear ordinal scale—ie, the clinical importance of a 1-point change varies according to the starting score. These factors limit the statistical approaches for the analysis of EDSS data and complicate interpretation of changes in score over time. Beyond these well-known drawbacks, there is also a lack of standardization in the less acknowledged technical details used to estimate progression events and the time to progression from baseline^5^.

These details are not uniformly applied across different clinical trials. Notably, determining confirmed EDSS progression from longitudinal data requires an explicit definition of parameters. This includes defining the reference baseline value, specifying criteria for progression (changes in EDSS according to the baseline value reported with variations across trials), specify rules for visits that are close to relapses and determining the time window for defining duration variables (e.g., time needed to confirm progression, typically reported as 3 or 6 months without an explicit definition of the tolerance window).

These technical details are crucial, as different definitions impact the number of events collected in a trial, influencing the study’s power to detect a significant treatment effect. Without a standardized approach to the primary outcome assessment, comparing trial results becomes challenging. This challenge is even greater for observational studies conducted in less strictly managed environments, where MS investigators often use self-made scripts, leading to unreliable and error-prone practices.

Recently, a new outcome, “progression independent of relapse activity” (PIRA), has been introduced for clinical trials in MS research^6^. The determination of PIRA events involves a growing number of parameters, making study results dependent on specific choices and necessitating scrupulous sensitivity analyses. Although a consensus paper for a uniform definition of PIRA was published^7^, shared definitions lack the granularity needed for practical calculation of PIRA events. The whole picture is even more complex when a composite measure of disability is used, involving other scales like frequently done including the Nine-Hole Peg Test (NHPT), the Timed 25-Foot Walk (T25FW) or the Symbol Digit Modalities Test (SDMT), requiring additional definitions, cut-off points and confirmation time windows.

The absence of a structured framework for progression computation criteria often results in a failure to exhaustively report the complete set of rules and cut-offs used, making it virtually impossible to precisely replicate study results. To address these challenges, we propose the adoption of a publicly available, automated, and customizable tool for computing MS progression outcomes. In a practical example, we demonstrate how subtle differences in parameter settings can alter the interpretation of trial results. This tool, integrated into an R package and designed for ease of use by clinicians, allows for standardized and explicit calculation of disability progression (EDSS based or composite) and PIRA.

## Methods

To demonstrate the effects of applying different parameter configurations, and to highlight the importance of reporting the complete set of criteria used, we re-analyzed data of the phase 3 placebo-controlled BRAVO clinical trial (ClinicalTrials.gov identifiers: NCT00605215), a study evaluating laquinimod vs placebo in relapsing-remitting MS patients^8^. The study was chosen since it had 12-week confirmed disability progression (CDP) as a secondary endpoint, based on EDSS changes, and it was negative on this endpoint with a p value of 0.06. This borderline result is well suited to show the impact on the trial result of changing the definition of CDP. The design and inclusion/exclusion criteria of the trial have been described elsewhere.^8^ Briefly, eligibility criteria included age 18–55 years, diagnosis of RRMS (revised McDonald criteria^9^, EDSS scores of 0–5.5, and presence of relapse activity in the previous 12–24 months.

We first tried to replicate the results of the BRAVO study on the CDP endpoint by using the criteria reported in the study and by varying all the non-reported parameters until we were able to fully replicate the results. We then repeated the same analysis by changing the base settings to create six alternative definitions of EDSS CDP, compatible with the definition reported in the published documentation (the paper and the trial protocol reported on clinicaltrial.gov) or according to common practices in MS clinical trials.

Following the indications provided in the published study, we defined CDP as an increase in the EDSS score of at least 1.0 point from baseline if the baseline score was between 0 and 5.0, or an increase of at least 0.5 points if the baseline score was at least 5.5; we detected CDP events using a fixed baseline and a confirmation interval of 12 weeks or more (confirmation happening in the visit subsequent the one with the event after at least 84 days and all the intermediate visits); the presence of relapses was not taken into account in the calculation of CDP. This set of criteria represented the starting point (Definition 0) that was the one replicating the trial results. We then designed the alternative scenarios (Definitions 1 to 4), as follows.

- Definition 1: patients who discontinued treatment early and had a progression occurring at the last available visit with no confirmatory visits were considered as progressed (as reported in the protocol of ORATORIO trial, ClinicalTrials.gov identifiers: NCT01194570).^10^ In the BRAVO trial^8^ it is not explicitly reported how an event on the last visit is considered.
- Definition 1*: all progressions occurring at the last available visit but lacking a confirmation visit were considered as events. In the BRAVO trial^8^ it is not explicitly reported how an event on the last visit is considered.
- Definition 2: the presence of relapses was taken into account in the calculation of progression. A visit cannot be used as baseline or confirmation if within 30 days from a relapse. This restriction was used in previous trials.^11^
- Definition2*: as the previous definition but excluding also the possibility to have an event in a time window of 30 days from a relapse.
- Definition3: progression events were confirmed at 12 weeks with a tolerance of 30 days on both sides.
- Definition4: a roving baseline scheme was adopted where a confirmed EDSS decrease determined a new baseline to compute future disability progression.^12^

To show the impact of using different definitions also on PIRA event calculations, we examined the time to first 12-week confirmed PIRA event as an alternative endpoint. We adopted the same base set of criteria used in Definition 0 to replicate the study results on CDP and required an absence of relapses in the 90 days before and the 30 days after the event and the confirmation visit for a progression to be classified as PIRA, as recommended in Müller et al.^7^ As before, we also studied three alternative scenarios.

- Definition P0: base settings as in Definition 0, and relapse-free intervals defining PIRA as in Müller et al.^7^
- Definition P1: patients who discontinued treatment early and had a progression occurring at the last available visit with no confirmatory visits were considered as progressed.^10^
- Definition P1*: all progressions occurring at the last available visit were included.
- Definition P2: progression events were confirmed at 12 weeks with a tolerance of 30 days on both sides.
- Definition P3: a roving baseline scheme was adopted where a confirmed EDSS decrease determined a new baseline to compute future disability progression^7,12^

For both endpoints, we used Cox proportional hazard models to compute hazard ratios (HRs) and their 95% confidence intervals (CI) by correcting for baseline EDSS score, log of the number of relapses in the previous 2 years, and country/geographical region for all parameter configurations^8^.

### A toolbox for MS outcome calculation: *msprog*

The *msprog* package (https://github.com/noemimontobbio/msprog) for R was developed as a tool for reproducible analysis of CDP in multiple sclerosis (MS) from longitudinal data. To simplify the use of our package and make it more accessible, a web application with a user-friendly graphical interface was also implemented and is available at the link https://msprog.shinyapps.io/msprog/. Its core function, *MSprog*, detects and characterizes the progression events of any clinical outcome measure we want to use to define disability progression (e.g. EDSS, NHPT, T25FW, SDMT, and so on). The data needed for calculation are the repeated assessments of the measures through time and the respective dates; and the date of relapses (if any). The function detects progression (or improvement) events sequentially by scanning the outcome values in chronological order, and classifies progression events as relapse-associated or relapse-independent based on their relative timing with respect to relapses. A number of qualitative and quantitative options for event detection are given as optional arguments that can be set by the user and reported as a complement to the results to ensure reproducibility. These include: the events to be detected (first or multiple, progression and/or improvement); the baseline scheme (fixed or roving); the minimum shift in the outcome measure corresponding to a valid change from the reference value; the length of the event confirmation period(s) with the relative tolerance; the minimum distance from the last relapse for a visit to be a valid baseline, event, or confirmation; the minimum interval for which a confirmed change must be sustained to be retained as an event; whether or not to include progressions occurring at the last visit; customizable relapse-free intervals around baseline visit, event, and confirmation visit to define PIRA; whether or not to re-baseline after relapses when searching for PIRA events. The amount and type of information to be included in the results table and to be printed out by the function can be set by the user as well. This allows, when desired, to keep track of the event detection process and ensure that the chosen criteria produce the expected output. Supplementary Table 1 lists the function arguments, detailing the role and default behavior of each of them.

## Results

### Summary of clinical trial results

The complete baseline demographic and clinical characteristics of the patients involved in the BRAVO trial were outlined in the trial report^8^ and can be found in Supplementary Table 2. For the purposes of this study, we focused our analysis solely on the patients who were part of the placebo arm (N=450) and those in the laquinimod arm (N=434). Patients treated with interferonβ-1a IM were excluded from the analysis (N=447).

According to the trial report, in our re-analysis of the study, laquinimod demonstrated a 31% reduction in 12-week CDP compared to the placebo (hazard ratio (HR) = 0.69, 95% confidence interval: 0.46–1.02, p = 0.06).

### Changing the settings with *msprog*

#### Confirmed Disability Progression

HRs quantifying the treatment effect on EDSS CDP, as measured according to the various definitions estimated by the msprog toolbox, are presented in Table 1 and Figure 1.

**Table 1.** Treatment effect on 12-week confirmed disability progression (CDP) under various scenarios.

|  | CDP definitions | n | Laquinimod | Placebo | HR (95% CI) | p-value |
| --- | --- | --- | --- | --- | --- | --- |
| Def 0 | Replication of published results. | 102 | 42 | 60 | 0.69 (0.46-1.02) | 0.06 |
| Def 1 | Progression events at last available visit evaluated as progression in patients with early treatment discontinuation with no confirmatory visit. | 117 | 44 | 73 | 0.59 (0.41-0.86) | 0.006 |
| Def 1* | Progression events evaluated as progression when occurring at the last available visit. | 141 | 54 | 87 | 0.60 (0.43-0.84) | 0.003 |
| Def 2 | A visit cannot be used as baseline or confirmation if within 30 days from a relapse | 98 | 41 | 57 | 0.72 (0.48-1.07) | 0.107 |
| Def 2* | A visit within 30 days from a relapse cannot be a valid baseline, event, or confirmation. | 86 | 33 | 53 | 0.63 (0.41-0.97) | 0.038 |
| Def 3 | Progression events confirmed at 12 weeks with a tolerance of 30 days on both sides | 113 | 48 | 65 | 0.72 (0.50-1.05) | 0.092 |
| Def 4 | Adoption of a roving baseline scheme where a confirmed EDSS decrease determines a new baseline to compute future disability progression. | 114 | 48 | 66 | 0.71 (0.49-1.04) | 0.078 |

Abbreviations: EDSS= Expanded disability Status Scale; CDP= Confirmed Disability Progression; Def=Definition; HR=Hazard ratio; 95%CI=95% Confidence Interval; n=number of events msprog parameters: Outcome="edss", conf\_weeks=12, event="firstprog"

**Figure 1.**
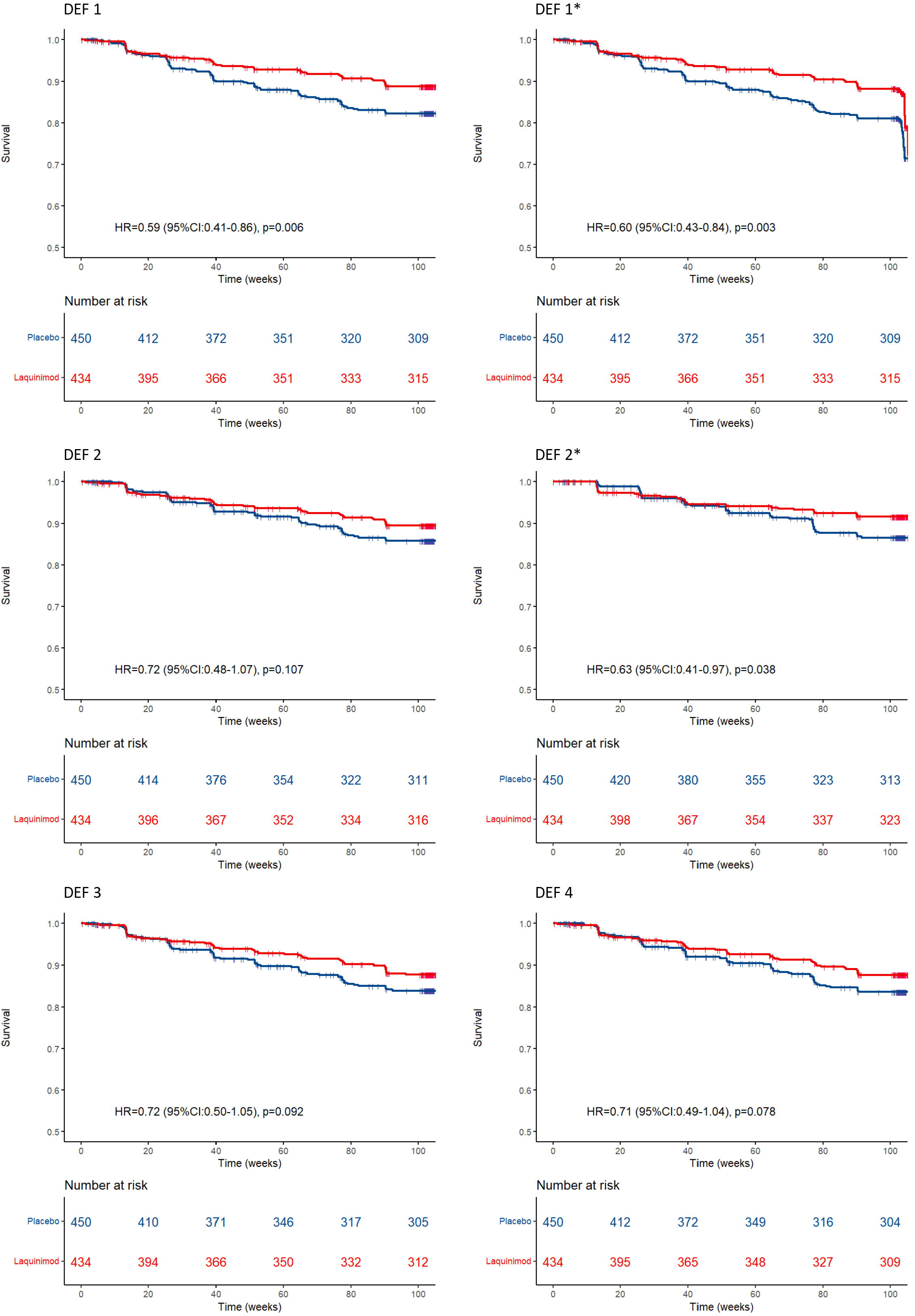
12-week EDSS confirmed disease progression (CDP) as identified by using different CDP definition. Def 1: progression event evaluated as progression in patients with early treatment discontinuation with no confirmatory visit; Def 1*: progression event evaluated as progression when occurring at the last available visit; Def 2: a visit cannot be used as baseline or confirmation if within 30 days from a relapse; Def 2*: a visit within 30 days from a relapse cannot be a valid baseline, event, or confirmation; Def 3: progression events were confirmed at 12 weeks with a tolerance of 30 days on both sides; Def 4: a roving baseline scheme was adopted where a confirmed EDSS decrease determined a new baseline to compute future disability progression

The treatment effect varied considerably depending on the definitions examined, with HRs fluctuating between 0.59 (risk reduction on CDP of 41%) and 0.72 (risk reduction on CDP of 28%). Also the p values, depending both on the effect size and on the number of events, fluctuated between 0.003 and 0.11, leading to different conclusion about the positivity of trial results. The most impactful change from the reference definition was the inclusion as an event of the progression that occurred to patients who discontinued treatment early and had a progression occurring at the last available visit with no confirmatory visits (Definition 1). That increased both the number of events and the treatment effect size (the HR passed from 0.69 [p=0.06] to 0.59 with a p=0.01). This definition was previously used in progressive clinical trials^10^. On the other hand, when using a symmetric tolerance interval of 30 days for 12-week confirmation visits (Definition 3), we obtained a smaller treatment effect on CDP as compared to the effect obtained with the definition used in the trial (the HR passed from 0.69, p=0.06 to 0.72, p=0.09) even if the number of events did not change considerably.

### PIRA

Table 2 showcases HRs illustrating the treatment impact on CDP using PIRA events, stemming from different definitions implemented through the use of the msprog toolbox. Using the same CDP definition as in the original paper, and defining the relapse-free intervals as in [Müller JAMA Neurol 2023] (Definition P0), we observed a treatment effect of 35% on PIRA (HR=0.65, 95% CI: 0.40-1.04, p=0.070). Under Definition P1, which includes progressions occurring at the last available visit for patients discontinuing treatment, a significant reduction in CDP is observed with an HR of 0.62 (95% CI: 0.39-0.97, p=0.036). Similarly, Definition P2, confirming progression events at 12 weeks with a tolerance of 30 days, shows an HR of 0.63 (95%CI: 0.40-1.01, p=0.054). Definition P3, employing a roving baseline scheme, detected a similar effect of treatment on PIRA (HR=0.65, 95%CI: 0.42-1.01, p=0.057).

**Table 2.** Treatment effect on 12-week confirmed progression independent on relapse activity (PIRA) under various scenarios.

|  | CDP definitions | n | Laquinimod | Placebo | HR (95% CI) | p-value |
| --- | --- | --- | --- | --- | --- | --- |
| Def P0 | Base settings as in Definition 0, and relapse-free intervals defining PIRA as in Müller et al.<sup>7</sup> | 74 | 29 | 45 | 0.65 (0.40-1.04) | 0.070 |
| Def P1 | Progression events at last available visit evaluated as progression in patients with early treatment discontinuation with no confirmatory visit. | 81 | 31 | 50 | 0.62 (0.39-0.97) | 0.036 |
| Def P1* | Progression events evaluated as progression when occurring at the last available visit. | 98 | 38 | 60 | 0.62 (0.41-0.93) | 0.021 |
| Def P2 | Progression events confirmed at 12 weeks with a tolerance of 30 days on both sides. | 75 | 29 | 46 | 0.63 (0.40-1.01) | 0.054 |
| Def P3 | Adoption of a roving baseline scheme where a confirmed EDSS decrease determines a new baseline to compute future disability progression. | 84 | 33 | 51 | 0.65 (0.42-1.01) | 0.057 |

Abbreviations: EDSS= Expanded disability Status Scale; CDP= Confirmed Disability Progression; Def=Definition; HR=Hazard ratio; 95%CI=95% Confidence Interval; n=number of events msprog parameters: Outcome="edss", conf\_weeks=12, event=" firstPIRA", relapse\_to\_bl=30, relapse\_to\_conf=30

## Discussion

Measuring disability and CDP in MS studies presents numerous challenges, which have been thoroughly explored and documented. Another contributing factor to the heterogeneity in the assessment of treatment effects on CDP is the absence of a standardized definition for estimating CDP events. This lack of uniformity is further compounded by incomplete reporting of all the necessary details essential for a unique assessment of CDP events. These details are in fact very different across clinical trials, and we showed here that fluctuations of such details can have a significant impact on the trial results, leading to different conclusions.

We’ve created a tool here to calculate CDP based on various definitions, that can be expanded further as required. The goal is to provide an open-access tool with a user-friendly interface and standardized output. This allows researchers to work with consistent definitions, reducing heterogeneity in CDP estimation. It will be essential to compare our software with tools developed by other researchers worldwide to enhance its accuracy. This is feasible because the software is freely available and programmed in R, the most widely used free language for statistical programming.

## Data Availability

Data used in this work were collected within the International Progressive MS Alliance project (IPMSA, award reference number PA-1603-08175). Access requests should be forwarded to the relevant data controllers.

## Acknowledgements

FB received the 2022 Biostatistic/Informatics Junior Faculty Award (grant code BI-2107-38160) awarded by the National MS Society.

NM received funding from the European Union – Next Generation EU – PNRR – MUR – M4C2 Investimento 1.3 - Public notice “Partenariati Estesi” - MNESYS – A multiscale integrated approach to the study of the nervous system in health and disease – CUP D33C22001340002.

