## Supplemental material for "Creating an automated tool for a consistent and repeatable evaluation of disability progression in clinical studies for Multiple Sclerosis"

**‘msprog’ package description**

The *msprog* package is publicly available on GitHub (https://github.com/noemimontobbio/msprog). Installation instructions are provided on the web page. The package includes a main function, *MSprog*, as well as some ancillary functions and two toy datasets for function testing.

The *MSprog* function detects and characterizes the progression events of a clinical outcome. It takes as input a longitudinal dataset containing the dates of the assessments and the relative outcome values, and (optionally) an additional dataset containing the dates of acute episodes. The artificial datasets *toydata_visits* and *toydata_relapses*, included in the package, provide an example of correctly specified input data. The other arguments (see Supplementary Table 1) specify qualitative and quantitative options to define and detect progression events, and to control the amount and type of information returned. The function prints out progress information, and returns a tabular description of all patients’ disease progression.

The package also contains the following ancillary functions:

- *compute_delta*: it computes the minimum shift corresponding to a valid change from the provided baseline value. It supports EDSS, NHPT, T25FW, and SDMT as outcome measure. The *compute_delta* function is used as default *delta_fun* in MSprog. Alternatively, a custom function can be provided by the user to support different outcome measures, or to specify different rules for the supported ones.
- *relapse_indep_from_bounds*: it can be used to specify custom relapse-free intervals to define PIRA. It takes left and right bounds around baseline, event and confirmation, and organizes them into a valid *relapse_indep* argument for function MSprog.

For further details about function usage and behavior, please refer to the package documentation (by typing, e.g., ‘?MSprog’). Detailed tutorials on usage and best practices are available as package vignettes, that can be displayed by typing *browseVignettes(‘msprog’)*.

**Supplementary Table 1. MSprog function arguments with relative description and default value (for optional arguments).**

| **Argument** | **Description** | **Default value** |
| --- | --- | --- |
| data | A data.frame containing longitudinal data, including: subject ID, outcome value, date of visit. | - |
| subj_col | Name of data column with subject ID. | - |
| value_col | Name of data column with outcome value. | - |
| date_col | Name of data column with date of visit. | - |
| outcome | Specifies the outcome type. Must be one of: 'edss', 'nhpt', 't25fw', 'sdmt', NULL (only accepted when specifying a custom delta_fun, see below). | - |
| subjects | Subset of subjects (list of IDs). If none is specified, all subjects listed in data are included. | NULL |
| relapse | (optional) data.frame containing longitudinal data, including: subject ID and relapse date. | NULL |
| rsubj_col | Name of subject ID column for relapse data, if different from outcome data. | NULL |
| rdate_col | Name of date column for relapse data, if different from outcome data. | NULL |
| delta_fun | Custom function specifying the minimum shift corresponding to a valid change from the provided baseline value. If none is specified (default), the following default criteria are used:   - EDSS: 1.5 points if reference is 0.0; 1.0 point if reference is between 0.5 and 5.0 (included); 0.5 points if reference is higher than 5.0. - NHPT: 20% of reference. - T25FW: 20% of reference. - SDMT: either 3 points or 10% of reference. | NULL |
| conf_weeks | Period before confirmation (weeks). | 12 |
| conf_tol_days | Tolerance window for confirmation visit (days). It can be an integer (same tolerance on left and right) or list-like of length 2 (different tolerance on left and right). In all cases, the right end of the interval is ignored if conf_unbounded_right is set to TRUE. | 30 |
| conf_unbounded_right | If TRUE, the confirmation window is unbounded on the right (e.g., “12 weeks or more”). | FALSE |
| require_sust_weeks | Minimum number of weeks for which a change must be sustained to be retained as an event (in addition to confirmation). | 0 |
| relapse_to_bl | Minimum distance from last relapse (days) for a visit to be a valid baseline (otherwise the next available visit is used as baseline). | 30 |
| relapse_to_event | Minimum distance from last relapse (days) for an event to be considered as such. | 0 |
| relapse_to_conf | Minimum distance from last relapse (days) for a visit to be a valid confirmation visit. | 30 |
| relapse_assoc | Maximum distance from last relapse (days) for a progression event to be considered as RAW. | 90 |
| event | Specifies which events to detect. Must be one of the following:   - 'firstprog' (first progression); - 'first' (only the very first event – improvement or progression); - 'firsteach' (first improvement and first progression – in chronological order); - 'firstprogtype' (first progression of each kind – PIRA, RAW, and undefined, in chronological order); - 'firstPIRA' (first PIRA); - 'firstRAW' (first RAW); - 'multiple' (all events, in chronological order). | 'firstprog' |
| baseline | Specifies the baseline scheme. Must be one of the following:   - 'fixed' (first valid outcome value); - 'roving_impr' (updated after each confirmed improvement; suitable in a first-progression setting to discard baseline fluctuations). - 'roving' (updated after each confirmed event; suitable in a multiple-event setting). | 'fixed' |
| relapse_indep | Specifies relapse-free intervals for PIRA definition. Must be given in the form produced by function  relapse_indep_from_bounds(b0, b1, e0, e1, c0, c1)  by specifying the intervals around baseline (b0 and b1), event (e0 and e1), and confirmation (c0 and c1). If none is specified, the following definition is used as default:  relapse_indep_from_bounds(0, 0, 90, 30, 90, 30)  i.e., absence of relapses in the 90 days before and 30 days after the event and the confirmation visit [Müller JAMA Neurol 2023]. | NULL |
| sub_threshold | If TRUE – and only if baseline is 'roving' or 'roving_impr' – move roving baseline at any sub-threshold confirmed event (i.e., any confirmed change in outcome measure, regardless of delta_fun). | FALSE |
| relapse_rebl | If TRUE, re-baseline after every relapse to search for PIRA events. | FALSE |
| min_value | Only consider progressions events where the outcome is >= a predefined value. | 0 |
| prog_last_visit | If TRUE, include progressions occurring at last visit (i.e. with no confirmation). If a numeric value N is passed, unconfirmed events are oncluded only if occurring within N weeks of follow up. | FALSE |
| include_dates | If TRUE, report date of event in results table. | FALSE |
| include_value | If TRUE, report value of outcome at event in results table. | FALSE |
| include_stable | If TRUE, subjects with no events are included in results table, with time2event = total follow up. | TRUE |
| verbose | One of:   - 0 (print no info); - 1 (print concise info after computation is complete); - 2 (print extended progress info for each subject). | 1 |

RAW: Relapse-Associated Worsening; PIRA: Progression Independent of Relapse Activity; EDSS: Expanded Disability Status Scale; NHPT: Nine-Hole Peg Test; T25FW: Timed 25-Foot Walk; SDMT: Symbol Digit Modalities Test.

**Supplementary Table 2. Baseline characteristics of the analyzed population enrolled in BRAVO RCT.**

|  | **Laquinimod (*n* = 434)** | **Placebo (*n* = 450)** |
| --- | --- | --- |
| Women, *n* (%) | 282 (65.0 %) | 321 (71.3 %) |
| Age (years) | | |
| Median (P25, P75) | 36.7 (29.6, 44.0) | 37.5 (30.3, 45.4) |
| Time from first MS symptom year | | |
| Median (P25, P75) | 4.9 (2.2, 9.3) | 4.7 (2.0, 9.7) |
| Time from MS diagnosis years | | |
| Median (P25, P75) | 1.2 (0.3, 3.8) | 1.2 (0.3, 4.0) |
| Patients with > 1 relapse in 1 year before entry, *n* (%) | 425 (97.9) | 435 (96.7) |
| Relapses in the previous year | | |
| Median (P25, P75) | 1.0 (1.0, 2.0) | 1.0 (1.0, 2.0) |
| Relapses in previous 2 years | | |
| Median (P25, P75) | 2.0 (1.0, 2.0) | 2.0 (1.0, 2.0) |
| EDSS score | | |
| Median (P25, P75) | 2.5 (1.5, 3.5) | 2.5 (1.5, 3.5) |
| Prior disease-modifying treatment for MS^a^, *n* (%) | 30 (6.9) | 27 (6.0) |
| % of patients with GdE lesions | 39.6 | 33.4 |
| Volume of T2 lesions (cm^3^) | | |
| Median (P25, P75) | 6.3 (2.3, 13.5) | 4.7 (1.7, 10.3) |
| Normalized brain volume (cm^3^) | | |
| Mean (SD) | 1,582 (96) | 1,586 (93) |

EDSS: Expanded Disability Status Scale; MRI: magnetic resonance imaging; GdE: gadolinium-enhancing; P25:25th percentile; P75: 75th percentile

^a^At any time before study entry. DMAMS included mitoxantrone, immunoglobulin (Ig), IgG, glatiramer acetate, IFNβ drugs, meglumine acridonacetate, and azathioprine
